# HCDPD: A Heterogeneous Causal Framework for Disease Pattern Detection in Medical Imaging

**DOI:** 10.1101/2025.04.15.25325904

**Authors:** Rongjie Liu, Chengchun Shi, Rui Song, Marc Niethammer, Tengfei Li, Hongtu Zhu

## Abstract

Understanding the causal effects of diseases on body organs through medical imaging is crucial for advancing research and improving clinical outcomes. This paper introduces a novel causal inference framework, Heterogeneous Causal Disease Pattern Detection (HCDPD), designed to map the complex causal pathways from early-stage diseases to latent disease patterns and their manifestation in organs as observed in later-stage medical images. HCDPD serves as a potential outcome framework for multivariate responses. It is particularly valuable in scenarios where patients exhibit significant heterogeneity, while normal controls remain relatively homogeneous. Through the application of advanced Bayesian inference techniques, our method effectively estimates both direct and indirect causal effects within the HCDPD framework. We applied HCDPD to the Osteoarthritis Initiative (OAI) dataset, successfully identifying and delineating diverse disease patterns across different patients. This capability provides critical insights that can inform early interventions and tailor personalized treatment strategies in clinical practice.

## 1 Introduction

Medical imaging techniques, such as Magnetic Resonance Imaging (MRI), are essential tools for examining the anatomy, functionality, and molecular pathways of various organs. These imaging methods significantly enhance the detection and analysis of biomarkers in both clinical and research settings (Oei et al. 2022, Kavur et al. 2021, Zhu et al. 2023). Imaging biomarkers are particularly critical for diagnosing conditions like dementia and for evaluating cardiovascular risks through cardiac imaging. This supports clinical decision-making, treatment planning, and the assessment of new therapies in clinical trials (Therriault et al. 2024).

Over time, the field has expanded to include a variety of methodologies aimed at detecting disease patterns. These range from functional regression and latent variable models to advanced image analysis techniques that leverage region-specific features to identify disease signatures (Liu & Zhu 2021, Huang et al. 2022, 2015, Davatzikos et al. 2008). Furthermore, the advent of deep learning has substantially enhanced automated feature extraction, facilitating the accurate identification of complex conditions such as brain tumors (Sharif et al. 2020, Manhas et al. 2022). Despite these advances, integrating causal inference with disease pattern detection within a potential outcome framework remains under-explored, highlighting a significant research gap.

The potential outcome framework evaluates outcomes under an intervention relative to hypothetical scenarios in which the intervention did not occur. Central to this framework are the concepts of potential or counterfactual outcomes, grounded in assumptions such as the Stable Unit Treatment Value Assumption (SUTVA), ignorability, and positivity. Comprehensive reviews of these concepts can be found in Rubin (2005) and Li, Ding & Mealli (2023). Recently, this framework has been extended to accommodate complex data types, such as spatial data, offering innovative methodologies and insights for causal inference across diverse research fields (Reich et al. 2021, Luo et al. 2024).

Our proposed method, the Heterogeneous Causal Disease Pattern Detection (HCDPD), introduces a novel potential outcome framework tailored for comparing high-dimensional medical images under different treatment conditions across the same subjects (Figure 1(a)). The data consists of observations {(**x**_*i*_, *g*_*i*_, *Y*_*i*_) : *i* = 1, …, *n*} collected from *n* independent subjects. Here, **x**_*i*_ denotes baseline covariates (e.g., age, gender), *g*_*i*_ represents an early-stage clinical diagnosis (with *g*_*i*_ = 0 for control subjects and *g*_*i*_ *>* 0 indicating case subjects), and *Y*_*i*_ = {*Y*_*i,s*_ : *s* ∈ 𝒮} is the observed high-dimensional medical image for subject *i*, where *s* indicates a pixel within a common spatial domain 𝒮.

**Figure 1:**
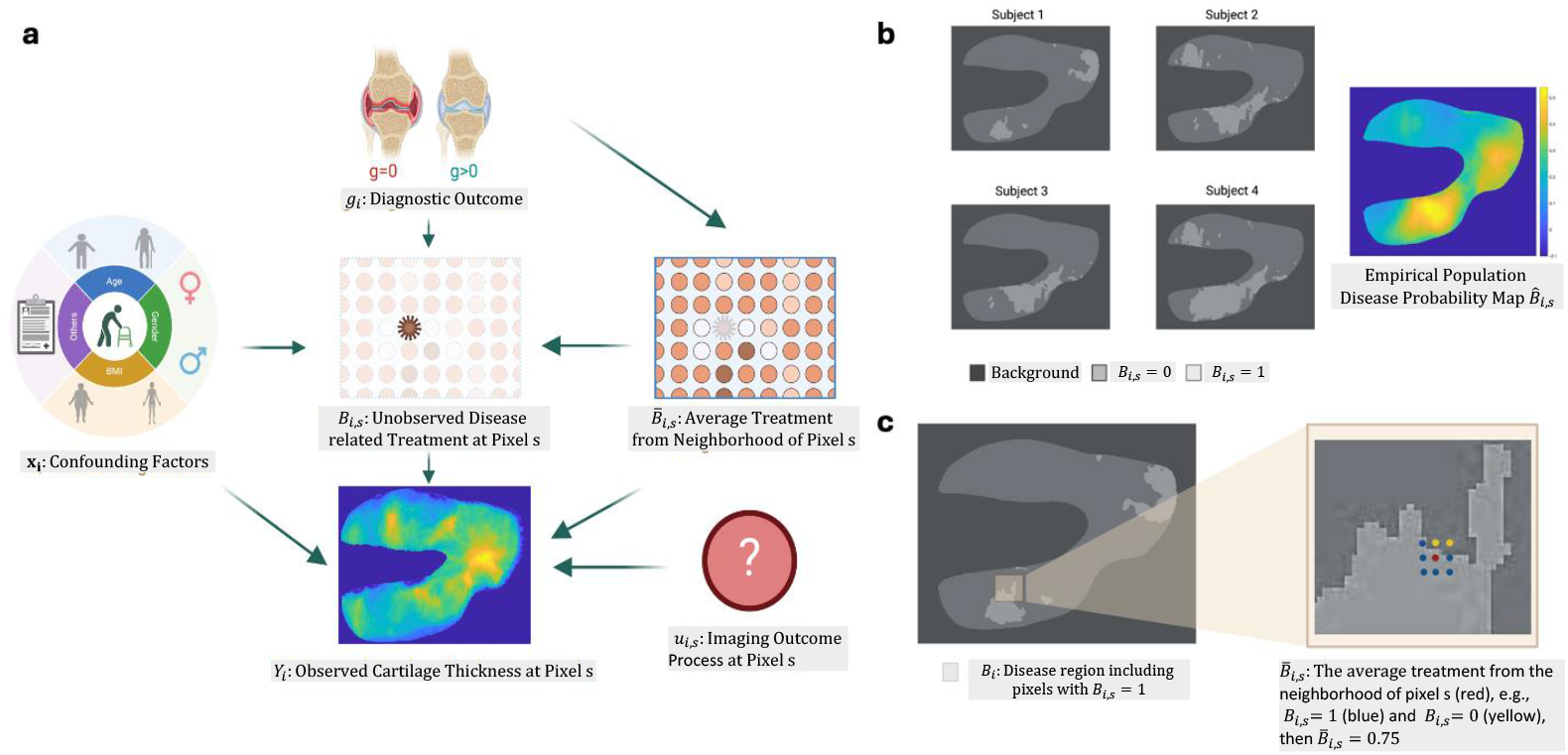
(a) The HCDPD framework: **x**_*i*_ represents baseline covariates (e.g., age, gender), *g*_*i*_ denotes an early-stage clinical diagnosis (*g*_*i*_ = 0: control, *g*_*i*_ *>* 0: case subjects), *Y*_*i*_ is a subsequent medical image, and *B*_*i,s*_ indicates the latent disease pattern. Causal inference challenges include: (b) spatial location heterogeneity among patients and (c) spatial spillover effects within each patient.

Under the classical causal inference framework, each individual *i* possesses multiple potential outcomes: *Y*_*i*_(0) and *Y*_*i*_(*g*_*i*_) for cases where *g*_*i*_ *>* 0, where *Y*_*i*_(*g*) = {*Y*_*i,s*_(*g*) : *s* ∈ 𝒮} represents the medical image under treatment condition *g*. The Individualized Treatment Effect (ITE) for individual *i* is defined as ITE_*i*_(*g*_*i*_) = *Y*_*i*_(*g*_*i*_) − *Y*_*i*_(0) for *g*_*i*_ *>* 0, while the Conditional Average Treatment Effect (CATE) is given by CATE_*i*_(*g*, **x**) = 𝔼 [*Y*_*i*_(*g*) − *Y*_*i*_(0)|**x**_*i*_ = **x**] = *µ*_*g*_(**x**) − *µ*_0_(**x**), with *µ*_*g*_(**x**) = 𝔼 [*Y*_*i*_(*g*)|**x**_*i*_ = **x**] for all *g* ≥ 0. In contrast to many existing approaches within the potential outcomes framework (Imbens & Rubin 2015, Rubin 2005, Li, Ding & Mealli 2023), significant challenges arise when aggregating ITE and CATE across individuals. We highlight two primary challenges in detail below.

The first challenge is **spatial location heterogeneity**, referring to the significant variability of disease-affected regions across different patients (Huang et al. 2015, Liu & Zhu 2021, Banerjee et al. 2017, Sharif et al. 2020, Shaker et al. 2017). To characterize this heterogeneity, we define an individual latent disease map for subject *i* as *B*_*i*_ = {*B*_*i,s*_ : *s* ∈ 𝒮}, where *B*_*i,s*_ = 1 indicates a disease-affected pixel and *B*_*i,s*_ = 0 denotes a normal pixel.

Disease-affected regions, denoted as 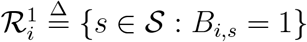, generally differ in size, shape, and spatial location among patients (Huang et al. 2015, Liu & Zhu 2021). Traditional approaches within the potential outcomes framework often neglect this individual-level spatial heterogeneity, failing to distinguish treatment effects between disease-affected pixels 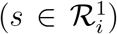 and unaffected pixels 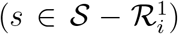. To demonstrate this issue, averaging the individual disease maps *B*_*i*_ across subjects results in an empirical population disease probability map (PDPM), as illustrated in Figure 1(b). The PDPM from real data shows regions with disease probabilities significantly greater than zero yet consistently below or near 0.5, underscoring substantial spatial location heterogeneity across individuals.

The second challenge is the **spatial spillover effect**, which emerges due to strong spatial dependencies among neighboring pixels. In disease detection, the presence of disease in one location can influence surrounding tissues or areas, violating SUTVA. Violation of SUTVA due to spatial spillover effects leads to biased and inconsistent estimates, potentially causing incorrect causal inferences (Corrado & Fingleton 2012). Figure 1(c) demonstrates an estimated disease map 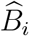, highlighting multiple detected disease-affected areas rather than isolated disease pixels. This observation suggests that a pixel’s disease status *B*_*i,s*_ is influenced by its neighboring pixels’ status, denoted as 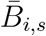, which we calculate by averaging the treatment statuses of adjacent pixels. Addressing this spatial spillover is critical for accurate and reliable causal assessments in spatial medical imaging analysis.

The literature on spatial causal inference can broadly be categorized into two main groups: methods that adjust for unmeasured confounders and methods that address spatial interference (Pollmann 2020, Reich et al. 2021). Within the first group, structured models such as Conditional Autoregressive (CAR) and Spatial Autoregressive (SAR) models are popular approaches for mitigating or eliminating confounding biases (Banerjee et al. 2003, Lee 2007). The second group, focusing on spatial interference, can be further divided into four distinct subcategories: (i) **Exposure Mapping**: Methods that define general forms of interference using exposure mapping (Aronow & Samii 2017). (ii) **Partial Interference**: Approaches that divide populations into non-overlapping blocks to manage intra-block interference (Sobel 2006, Tchetgen & VanderWeele 2012, Pollmann 2020). (iii) **Local or Network-based Interference**: Techniques employing local or network-based assumptions to evaluate exposure effects among experimental units situated in geographical spaces or networks (Bakshy et al. 2014, Puelz et al. 2022). (iv) **Congestion or Pricing Interference**: Recent advancements that introduce novel estimators to address interference arising from congestion or pricing dynamics in two-sided markets (Munro et al. 2021, Johari et al. 2022). Although these methodologies constitute a robust toolkit tailored to handle the diverse challenges posed by spatial data and interference patterns, none are directly applicable to the specific problem of disease pattern detection described earlier.

In this paper, we introduce HCDPD with four major contributions as follows: <u>(i)</u>. We are among the first to apply the potential outcomes framework to detect disease patterns in medical imaging, effectively quantifying the heterogeneous causal effects of disease on the human body. <u>(ii)</u>. We propose the HCDPD framework, which integrates a functional outcome model, a latent exposure model, and SAR priors. The functional outcome model accounts for the effects of baseline covariates and unobserved latent disease maps on imaging outcomes, while the latent exposure model estimates the probability of disease-affected pixels based on observed covariates. The SAR priors capture spatial spillover effects in both imaging outcomes and latent disease maps. <u>(iii)</u>. We utilize Markov Chain Monte Carlo (MCMC) techniques to quantify uncertainties in disease pattern detection and parameter estimation. This approach allows us to estimate posterior distributions, which facilitate causal inference regarding the impact of treatments or exposures on outcomes. (iv). We perform sensitivity analyses to evaluate the impact of different prior distributions or model specifications on the causal estimates. Additionally, we assess counterfactuals using the estimated parameters to infer causal effects of various interventions on different outcomes. The code and data corresponding to our proposed method are also publicly available on GitHub.

The paper is structured as follows: Section 2 outlines the motivation based on real Osteoarthritis Initiative (OAI) data and the key scientific questions. Section 3 introduces the HCDPD framework, including its model structures, Bayesian estimation methods, and a sensitivity analysis. Section 4 demonstrates the effectiveness of proposed method through simulations and experimental validation. Section 5 presents an analysis of real OAI data, followed by the conclusions. Finally, Section 6 provides a concise summary of the study.

## 2 Data Description

This paper analyzes data from the OAI, a comprehensive, multi-center observational study conducted over a span of more than ten years (2004-2019), involving both men and women aged from 45 to 79 (Attur et al. 2020). The study enrolled 4,796 participants between 2004 and 2006, consisting of three sub-cohorts: a progression sub-cohort (n=1,389), an incidence sub-cohort (n=3,285), and a normal control unexposed reference sub-cohort (n=122). Detailed information about the OAI is available at http://www.oai.ucsf.edu/. This study focuses primarily on baseline observations to explore the causality between individual OA patterns and corresponding knee cartilage thickness maps.

Knee OA is the leading cause of chronic pain, affecting approximately 21.2% of the U.S. population (Fallon 2023). It is a primary contributor to disability, often necessitating medical intervention. With an aging population and rising obesity rates, the prevalence of knee OA is projected to increase significantly (Heidari 2011). Previous research has established correlations between clinical variables such as age, gender, and body mass index (BMI) and the severity of knee OA (Schaefer et al. 2017, Huang et al. 2022). Furthermore, comorbid conditions like diabetes mellitus and obesity have been associated with increased pain severity in knee OA patients (Shin 2014). Thus, our study aims to identify individual-specific disease patterns of knee OA and evaluate their causal impacts.

MRI is a valuable tool for assessing knee joint degeneration due to its ability to capture detailed characteristics of the joint, including cartilage morphology and biochemical composition (Huang et al. 2022). This study mainly focuses on the left knee femoral cartilage (FC) thickness maps extracted from the MRI scans of 4,418 subjects (2,566 females and 1,852 males) at baseline in OAI. The detailed image preprocessing steps can be found in Section 5.1. The MR images, acquired using 3.0 Tesla Siemens Trio MRI scanners, have uniform dimensions of 384 × 384 × 160 with a resolution of 0.36 × 0.36 × 0.7 mm^3^ per voxel, and are labeled with Kellgren-Lawrence grades (KLG, Kellgren & Lawrence 1957) ranging from 0 (normal) to 4 (severe osteoarthritis). The detailed demographic and KLG information in our study are summarized in Table 1.

**Table 1:** The detailed demographic (Gender, BMI, and Age) and KLG information in our study. The mean and standard deviations for BMI and Age are reported for each KLG group as well.

|  | KLG=0 | KLG=1 | KLG=2 | KLG=3 | KLG=4 | Total |
| --- | --- | --- | --- | --- | --- | --- |
| Sample size | 1,752 | 793 | 1,120 | 613 | 140 | 4,418 |
| Gender (F/M) | 993/759 | 474/319 | 718/402 | 327/286 | 54/86 | 2,566/1,852 |
| BMI (kg/m<sup>2</sup>) | 27.2±4.5 | 28.5±4.5 | 29.7±4.9 | 30.4±4.9 | 29.6±3.8 | 28.6±4.8 |
| Age (years) | 59.3±9.2 | 61.2±9.1 | 61.6±8.8 | 64.2±8.8 | 65.7±8.4 | 61.1±9.2 |

This paper seeks to address three key scientific questions as follows: (Q1) How can we estimate ITEs for all subjects with *g*_*i*_ *>* 0? (Q2) How can we establish the causal pathway from KLG to individual OA-related abnormal patterns to corresponding knee cartilage thickness maps? (Q3) How can we evaluate the spatial location heterogeneity across patients? The subsequent sections of the paper present a causal inference framework designed to answer these questions.

## 3 Methodology

To address (Q1)-(Q3) in Section 2, we introduce the HCDPD framework, including causal estimands and identification assumptions (Section 3.1), statistical modeling (Section 3.2), and Bayesian inference (Section 3.3).

### 3.1 Causal Estimands and Identification Assumptions

<u>To address (Q1)</u>, our first set of causal estimands is all ITEs in the disease group. Recall that ITE_*i*_(*g*_*i*_) = (ITE_*i,s*_(*g*_*i*_) : *s* ∈ 𝒮) = *Y*_*i*_(*g*_*i*_) − *Y*_*i*_(0) for the *i*-th subject with *g*_*i*_ *>* 0. Since we only have *Y*_*i*_(*g*_*i*_) in the disease group, it remains to estimate *Y*_*i*_(0). We estimate *Y*_*i*_(0) via regression to learn its conditional mean 𝔼 [*Y*_*i*_(0)|**x**_*i*_]. This leads to the following ITE estimator:

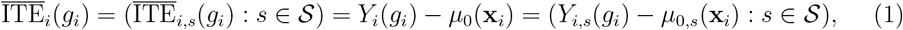

where *µ*_0_(**x**_*i*_) = (*µ*_0,*s*_(**x**_*i*_) : *s* ∈ 𝒮) = (*E*[*Y*_*i,s*_(0)|**x**_*i*_] : *s* ∈ 𝒮).

We begin by recalling and introducing some notations. For each *s* ∈ 𝒮, we denote its set of spatial neighboring grids (e.g., the nearest neighboring grids) in 𝒮 as 𝒩 (*s*). For the *i*-th subject with *g*_*i*_ *>* 0, recall that the disease region is defined by 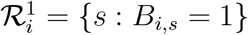. We define the neighboring closure of 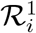 as 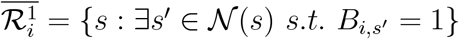. Furthermore, we define 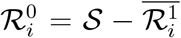 as the normal region of the *i*-th subject, which is an open set of 𝒮. If the *i*-th subject belongs to the control group, then 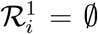 and 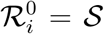. We next impose the following stability condition.

#### Assumption 3.1.

*(Stability): The distribution of* (*Y*_*i,s*_(*g*_*i*_), **x**_*i*_) *for each* 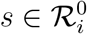 *in the i-th case subject is the same as that of* {(*Y*_*i*_*′*_,*s*_(0), **x**_*i*′_)} *for all i′ in the control group*.

This assumption requires that the organ within the normal region of a case subject remains unchanged. It is grounded in the relative stability of organ structure and function across normal subjects, given the covariates (e.g., age) in **x**. The stability assumption will play a key role in modeling treatments and imaging outcomes in Section 3.2.

We need an unconfounded assignment mechanism assumption such that the treatment assignment is independent of the potential outcomes conditional on **x**.

#### Assumption 3.2.

*(Unconfoundedness): The treatment assignment g*_*i*_ *is independent of the potential outcomes* {*Y*_*i*_(*g*) : *g* ≥ 0} *conditional on* **x**_*i*_.

Assumption 3.2 states that there is no unmeasured confounding.

#### Assumption 3.3.

*(Positivity):* 0 *< c*_min_ *< P* (**x**_*i*_|*g* = 0)*/P* (**x**_*i*_|*g*+) *< c*_max_ *<* ∞ *hold for all* **x**_*i*_ *in the g*+ *group, where P* (**x**|*g*) *denotes the conditional distribution of* **x** *in the* [*g*] *group*.

Assumption 3.3 is related to **overlap and balance**, referring to the similarity in the distributions of the covariates between the *g*+ and *g* = 0 groups. Under Assumptions 3.1 and 3.3, we can consistently estimate 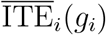 and then approximate the disease region 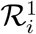 through modeling the effect of *B*_*i*_ = {*B*_*i,s*_ : *s* ∈ 𝒮} on 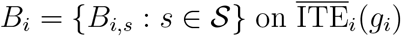 for each *i* in the [*g*+] group.

<u>To address (Q2) and (Q3)</u>, our first strategy is to construct an estimator of the underlying disease region *B*_*i*_ across all pixels *s* for the *i*-th patient. In practice, the *B*_*i*_ can be approximated by either frequentist or Bayesian approaches. More details will be discussed in Section 3.3. Next, to explicitly establish the causal pathway *g*_*i*_ → *B*_*i*_ → *Y*_*i*_ in Figure 1 (a), we also need to model *p*(*Y*_*i*_(*g*_*i*_)|*g*_*i*_, **x**_*i*_) as follows:

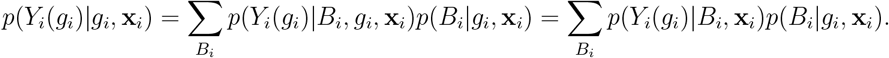

We need the following assumption about *p*(*Y*_*i*_(*g*_*i*_)|*B*_*i*_, **x**_*i*_).

#### Assumption 3.4.

*(Conditional Independence and Spatial Interference):*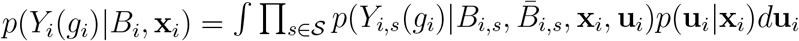, *where* **u**_*i*_ = (*u*_*i,s*_ : *s* ∈ 𝒮) *represents the spatial random effects*, 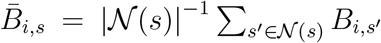 *is the average treatment from* 𝒩 (*s*), *and* | 𝒩 (*s*)| *is the number of pixels within* 𝒩 (*s*).

In Assumption 3.4, we address spatial interference specifically through information contained within 𝒩 (*s*) and introduce **u**_*i*_ to capture spatial correlations among *Y*_*i*_(*g*). These points will be elaborated in Section 3.2. Furthermore, to characterize OA-related spatial spillover effects from neighborhood 𝒩 (*s*), we incorporate the average treatment indicator 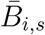 alongside *B*_*i,s*_ into the conditional distribution of *Y*_*i,s*_(*g*_*i*_). Similar strategies are commonly employed in models such as Gaussian hidden Markov models (Besag 1986) and Potts models (Krahenbühl & Koltun 2011). The primary function of 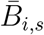 is to quantify the extent of local disease prevalence around pixel *s*. Practically, other statistics derived from *B*_*i,s*_ could also be employed to enhance modeling of spatial spillover effects.

To ensure the validity of our assumptions, we verify them using real data, detailed in the Supplementary Material S4. Under Assumptions 3.1-3.4, we introduce another set of causal estimands at pixel *s* for the *i*-th patient in the [*g*+] group. These estimands capture individual disease region patterns and potential spatial heterogeneity across patients. We define the total spatial effect (TSE) at pixel *s* as follows:

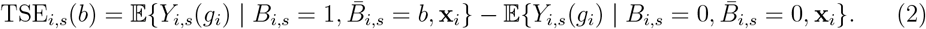

Furthermore, the TSE at pixel *s* can be divided in the direct spatial effect (DSE) and the indirect spatial effect (ISE) as follows:

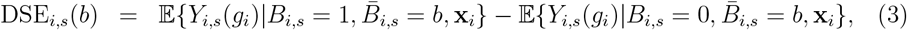

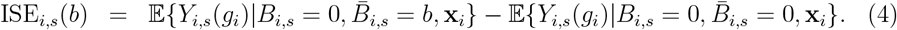

Given *B*_*i*_, the TSE captures the overall causal relationship between *B*_*i*_ and *Y*_*i*_. Importantly, the indirect spatial effect, ISE_*i,s*_(*b*), effectively models spatial spillover by quantifying how the surrounding disease region influences the cartilage thickness outcome at pixel *s*.

### 3.2 Modeling Treatment and Imaging Outcomes

To explicitly model *g*_*i*_ → *B*_*i*_ → *Y*_*i*_ in Figure 1, we introduce a hierarchical image-on-scalar regression model, including (i) a functional outcome model for 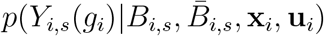 in Assumption 3.4 and (ii) a latent exposure model for 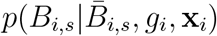.

#### (i) Functional outcome model

Given the OA-related treatment map, to characterize the effects of baseline covariates and treatment on imaging outcomes while accounting for spatial carryover effect and individual random variation, we introduce a functional outcome model as follows,

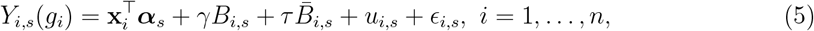

where ***α***_*s*_ is a *p*_*x*_-dimensional vector representing the fixed effects from observed confounding factors at pixel *s*. Parameters *γ* and *τ* quantify the OA-related treatment effects directly at pixel *s* and from its spatial neighborhood 𝒩 (*s*), respectively. We assume uniform OA-related treatment effects across pixels within the disease region 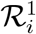. Additionally, the terms {*u*_*i,s*_, *s* ∈ 𝒮} represent individual stochastic imaging outcomes modeled as a zero-mean Gaussian process, and *ϵ*_*i,s*_ denotes Gaussian measurement error with distribution 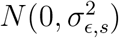. Thus, under model (5), the potential imaging outcome at pixel *s* primarily depends on the OA-related treatments at pixel *s* and its neighborhood, alongside effects from observed confounding factors. Figure 1(a) presents the Directed Acyclic Graph (DAG) illustrating this hierarchical image-on-scalar regression model.

In order to infer the individual processes *u*_*i,s*_ from the imaging outcomes, we employ the Bayesian functional principal component analysis approach (FPCA, Kowal & Bourgeois 2020, Zeng et al. 2021), where the observed imaging outcomes are projected into lower-dimensional representations and then the first few dominant principal components as the predictors in model (5). Specifically, we assume the individual imaging outcome process *u*_*i,s*_ has the following Karhunen-Loeve decomposition 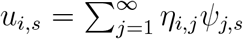, where 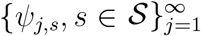 and 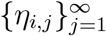 are, respectively, the corresponding normal orthogonal eigenfunctions and principal component (PC) scores of the *i*-th subject. In practice, we usually posit a similar model that truncates to the first *J* PCs of the imaging outcome process:

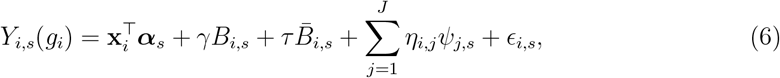

where the eigenfunctions 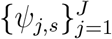 are derived based on the subjects at normal stage (Liu & Zhu 2021, Huang et al. 2022). Furthermore, we assume that the PC scores 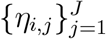 come from the zero-mean Gaussian distribution with shrinking variances, i.e., 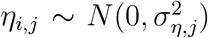 and 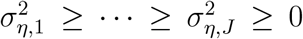. We select the truncation term *J* based on the fraction of explained variance (FEV) being greater than 85%, namely 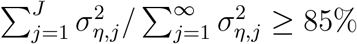.

#### (ii) Latent exposure model

To further characterize the latent OA-related treatment map, we propose a latent exposure model:

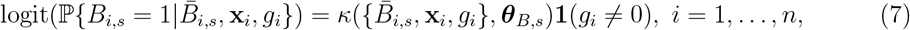

where logit(·) denotes the logit function and 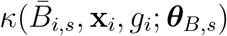 is a function parameterized by ***θ***_*B,s*_, representing the log odds ratio for case subjects at pixel *s*. For control subjects (*g*_*i*_ = 0), the log odds ratio is zero, implying *B*_*i,s*_ = 0 for all pixels, consistent with the assumption 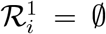. Typically, the function *κ*(·) is specified using domain knowledge. A common linear form is 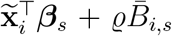, where 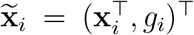 and ***θ***_*B,s*_ = (***β***_*s*_, *ϱ*) captures effects related to baseline covariates, KLG levels, and neighborhood influences. More complex structures, including nonlinear functions or higher-order interactions, can also be considered.

### 3.3 Posterior Inference of Causal Estimands

We first introduce a prior independence assumption, ensuring independent prior distributions for ***θ***_*Y*_ in model (5) and ***θ***_*B*_ = {***θ***_*B,s*_}_*s*∈𝒮_ in model (7):

#### Assumption 3.5.

*(Prior Independence): Parameters* ***θ***_*Y*_ *(imaging outcome model) and* ***θ***_*B*_ *(exposure assignment model) are distinct and independent a priori*.

Assumption 3.5, common in Bayesian causal inference, facilitates model specification and simplifies computational procedures (Li, Ding & Mealli 2023). Under Assumptions 3.1– 3.5, the posterior distribution of ***θ***_*Y*_ is independent of ***θ***_*B*_. Thus, given the latent disease regions 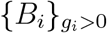 and priors *π*(***θ***_*Y*_), we fit the functional outcome model (5), sample from the posterior of 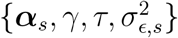, and construct FPCA representations for {*u*_*i,s*_}. Similarly, with the latent disease regions 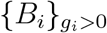 and prior *π*(***θ***_*B*_), we fit the latent exposure model (7) and sample from the posterior distribution of ***θ***_*B*_. We then derive the plug-in estimator *µ*_0_(**x**_*i*_) in (1) as 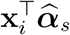. Consequently, we estimate the individual treatment effect (ITE) for each subject as 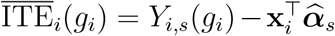 and the spatial causal estimands: 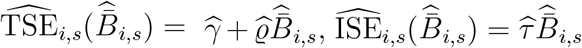 and 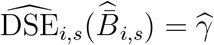. Note that the estimated DSE remains consistent across pixels and subjects.

The central aspect of estimating causal estimands is identifying the disease region *B*_*i*_ for each individual *i* ∈ [*g*+]. In a frequentist framework, we compute the standardized Individual Treatment Effect (SITE) estimator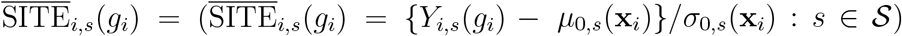, where 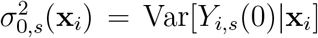. Under Assumptions 3.1–3.3, 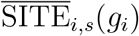 is consistently estimable without explicit distributional assumptions for *Y*_*i*_(*g*_*i*_).

Thus, we approximate the disease region as: 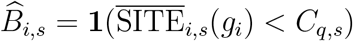 for each *i* ∈ [*g*+], where *C*_*q,s*_ is the *q*% quantile (commonly 0.5 or 0.1) of the reference distribution ℱ_0,*s*_, consistently estimated from the control group per Assumption 3.1. In contrast, the Bayesian approach provides both point estimates and posterior distributions of *B*_*i*_. Given posterior samples of ***θ***_*Y*_ and ***θ***_*B*_, we generate *M* samples 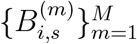 from the conditional posterior distribution. Then, we estimate the disease region as: 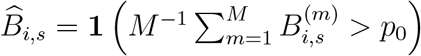 for each *i* ∈ [*g*+], where *p*_0_ ∈ (0, 1) is a predefined threshold. We also derive individual posterior disease probability maps as 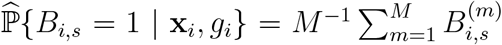 for each *i* ∈ [*g*+]. Additionally, the overall Population Disease Probability Map (PDPM) and subgroup-specific PDPMs based on KLG levels (*k* = 1, 2, 3, 4) are computed as 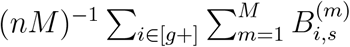 and 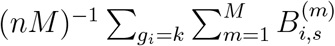, respectively.

To address uncertainty, correlation, and sparsity in spatial effects on imaging outcomes and OA-related latent exposures, we adopt a Bayesian approach utilizing spatial autoregressive spike-and-slab priors for parameters ***α***_*s*_ and coefficients in ***θ***_*B*_. Specifically, for the *l*-th element *α*_*l,s*_ in ***α***_*s*_, we impose the following prior:

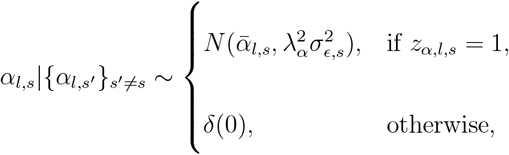

where 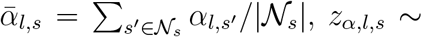 Bernoulli(*ν*_*α,l,s*_), and *δ*(0) denotes the degenerate distribution concentrated at zero. The complete details on priors and posterior inference are derived via Gibbs sampling, with specifics provided in the Supplementary Material S1 and S2.

A crucial aspect in disease region detection using MCMC sampling of *B*_*i*_ is selecting the threshold *p*_0_. The value of *p*_0_ significantly influences the extent of identified disease regions—smaller values result in larger detected regions, an ongoing challenge in anomaly detection research (Muñoz-Ramírez et al. 2022). Additionally, this threshold affects the estimation of causal estimands. We set *p*_0_ = 0.5 for simulations and real data analyses. To further examine the sensitivity of this choice, we will perform a detailed sensitivity analysis, exploring how various *p*_0_ values influence the variability of causal estimands.

## 4 Simulation Studies

We assessed the performance of the proposed framework through simulation studies using semi-realistic data derived from actual 2D femoral cartilage (FC) thickness maps of the left knee obtained from the OAI study. Detailed descriptions of the OAI dataset and image preprocessing procedures are provided in Section 5. In our simulation design, we incorporated key demographic and clinical covariates, Gender (coded as 1 for females), Age, and BMI, as potential confounding factors. In addition, we included two interaction terms: Gender*×*BMI and Gender*×*Age, to capture possible interaction effects. Both Age and BMI were standardized via Z-transformation. For both simulation studies and real data analyses, we employed consistent Gibbs sampling settings. Each MCMC run consisted of 2,200 iterations, with the first 200 iterations discarded as burn-in. To mitigate autocorrelation among samples, we implemented a thinning interval of 20, yielding 100 posterior samples. Posterior means computed from these thinned samples were used as point estimates for all model parameters. The convergence of our MCMC sampling was checked using multiple trace plots (see the Supplementary Material S3).

The true parameters for simulation settings were based on coefficients ***α***_*s*_ and 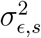 estimated from normal controls in the real dataset. Disease regions for each simulated patient were randomly selected across cartilage pixels, varying in size, shape (square or circle), and number. The disease-related coefficients were set as *γ* = −0.17 and *τ* = −0.22. Additionally, the first three principal components (PCs) from the real data analysis were used to generate individual stochastic imaging outcomes *u*_*i,s*_, with PC scores drawn from *η*_*i,j*_ ∼ *N* (0, 8 − 2*j*) for *j* = 1, 2, 3.

Given the generated disease regions and confounding factors, we fitted our latent exposure model (7), where 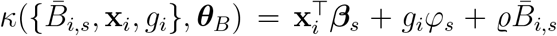, consistent with our real data analysis. The estimated coefficients ***β***_*s*_, *φ*_*s*_, and *ϱ* served as the true parameters for simulations. We generated 100 datasets, each containing 320 synthetic thickness maps (200 normal controls and 120 patients with disease). Example thickness maps and disease regions for five randomly selected subjects are displayed in Figure 2 (Columns 1 and 2).

**Figure 2:**
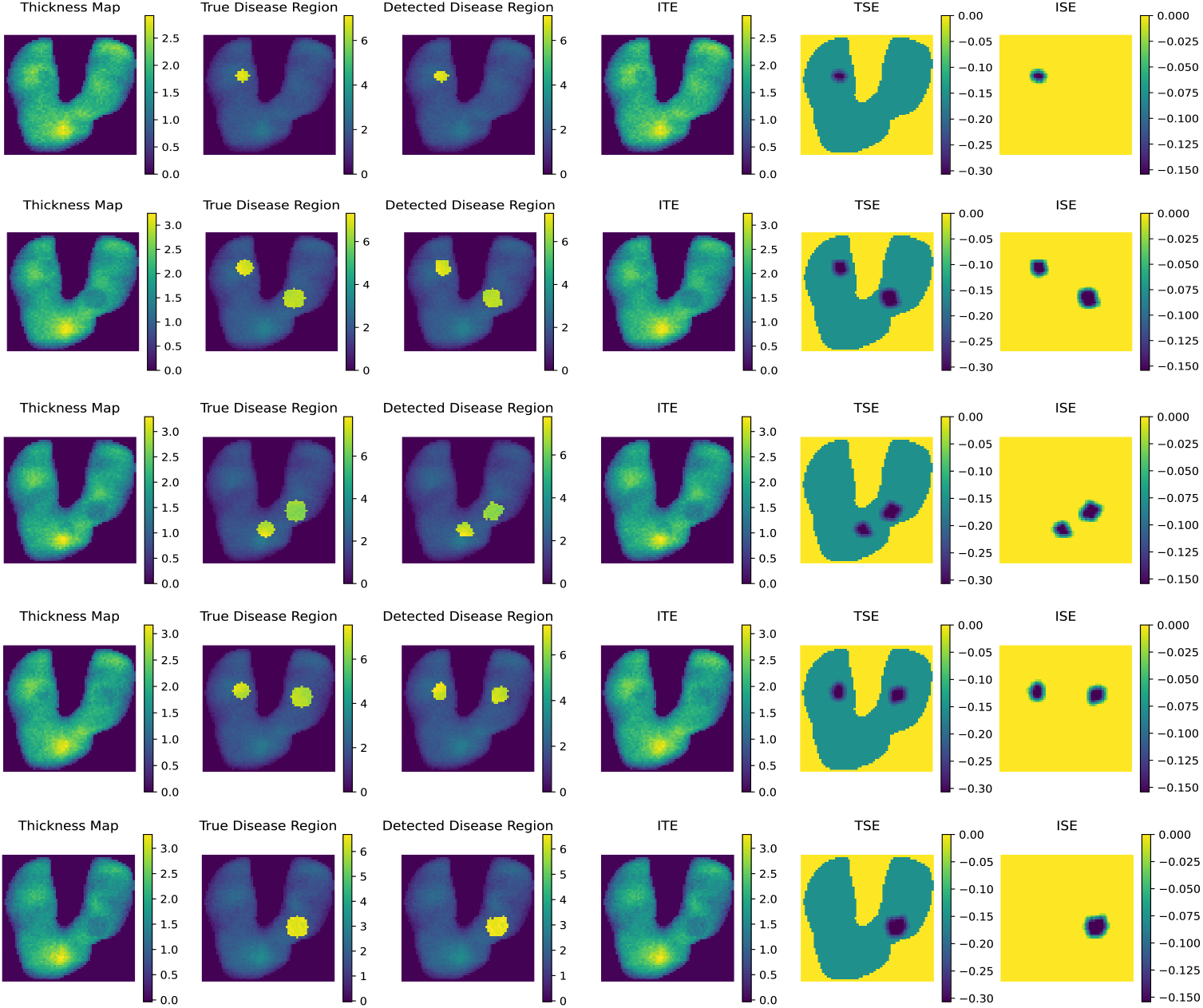
Simulated thickness maps (Column 1), true disease regions (Column 2), detected disease regions (Column 3), and estimated causal effects (ITE (Column 4), TSE (Column 5), and ISE (Column 6)) for five randomly selected subjects.

We applied our proposed method to these simulated datasets. The detected disease regions for selected subjects are shown in Figure 2 (Column 3). The overall accuracy was assessed using five metrics: Rand Index (RI), Adjusted Rand Index (ARI), Homogeneity (HOM), Completeness (COM), and Normalized Mutual Information (NMI). Averaged metrics per simulation are summarized in Figure S1. Our findings indicated: (i) high consistency between true and detected regions, with average RI around 0.95 and ARI around 0.80; (ii) HOM, COM, and NMI metrics consistently above 0.65, suggesting effective capture of disease region patterns. Additionally, the small variability across simulations (standard deviation *<* 0.015) demonstrated robustness in detecting disease regions.

We evaluated the estimation performance of the varying coefficients {***α***_*s*_, ***β***_*s*_, *φ*_*s*_}, and causal estimands (ITE, TSE, DSE, and ISE). For each simulation dataset, we calculated accuracy (ACC) in detecting sparsity structures in {***α***_*s*_, ***β***_*s*_, *φ*_*s*_} and mean squared errors (MSE) defined as:

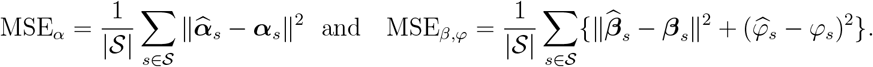

The results summarized in Figure S2 (left) showed average MSE and ACC values of approximately 0.6 and 0.7 for ***α***_*s*_, and 1.0 and 0.85 for {***β***_*s*_, *φ*_*s*_}, respectively, indicating robust performance in sparsity detection and parameter estimation.

Next, we examined causal estimands (ITE, TSE, DSE, ISE). Figure 2 (Columns 4-6) displays estimated ITE, TSE, and ISE for selected subjects, demonstrating causal effect patterns closely aligned with disease region patterns. The MSEs for TSE and ISE across all diseased patients were computed as:

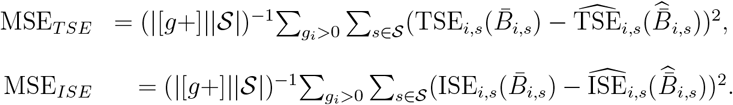

Figure S2 (right) summarizes these results, with DSE yielding an MSE of 0.0002, highlighting our method’s reliability in estimating causal effects.

Finally, we conducted two sensitivity analyses to evaluate robustness: (i) varying the threshold *p*_0_ in MCMC sampling and (ii) modifying Assumption 3.2 about unmeasured confounding. For the first analysis, we tested *p*_0_ values ranging from 0.3 to 0.8 and found negligible impacts on TSE and ISE estimation (Figure S3, left). In the second analysis, we introduced a binary unmeasured confounder *w*_*i,s*_, extending model (6) as:

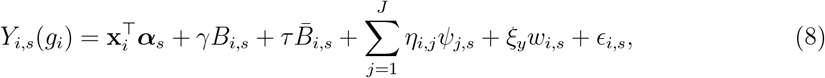

where *ξ*_*y*_ measures confounder influence. Testing *ξ*_*y*_ values from 0 to 0.15, we observed minimal bias for *ξ*_*y*_ ≤ 0.1, with biases increasing moderately thereafter (Figure S3, right). These analyses demonstrate our method’s robustness to moderate violations of Assumption 3.2.

## 5 Real Data Analysis

### 5.1 MRI data preprocessing

In this study, we implemented the image preprocessing pipeline introduced by Huang et al. (2022) to derive two-dimensional (2D) FC thickness maps from MRI data obtained in the OAI study. This pipeline comprises four main steps: segmentation and meshing, computation of 3D thickness maps, registration, and projection onto a 2D plane. First, we segmented the FC region from 3D MRI scans using a U-Net-based neural network (Huang et al. 2022). After segmentation, we constructed triangular meshes for each FC region using the marching cubes algorithm. Cartilage thickness at each mesh vertex was then calculated by measuring the shortest distance from the vertex to the opposite cartilage surface. Next, the cartilage meshes were spatially aligned to a common atlas space via a deep registration network. This step ensured consistency by mapping each MR image to an unbiased atlas, which was previously constructed using an atlas-building method (Huang et al. 2022). In the final stage, we projected the 3D atlas-aligned points onto a 2D plane and interpolated the thickness measurements to generate spatially coherent, flattened 2D FC thickness maps. Ultimately, we obtained baseline left-knee FC thickness maps for 4,418 subjects (2,566 females and 1,852 males) from the OAI study.

### 5.2 Addressing questions (Q1)-(Q3)

We fitted our proposed models (5) and (7) to the OAI data, in which 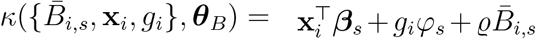. Besides the extracted left knee FC thickness maps, We also considered some baseline covariates (Age, Gender, BMI) and their interactions (Gender*×*BMI and Gender*×*Age) as possible confounders, where both BMI and Age were normalized through the Z-transformation. By applying the FPCA, we chose the top 30 PCs that can explain over 85% of variance in the imaging outcomes. The illustration of all the selected PCs can be found in the Figure S5. The estimated varying coefficients {***α***_*s*_, ***β***_*s*_, *φ*_*s*_} are presented in Figure S6. To more effectively assess the significance of the estimated effects and identify regions with notable impact, the t-test statistics for the varying coefficients across pixels are presented in Figure 3.

**Figure 3:**
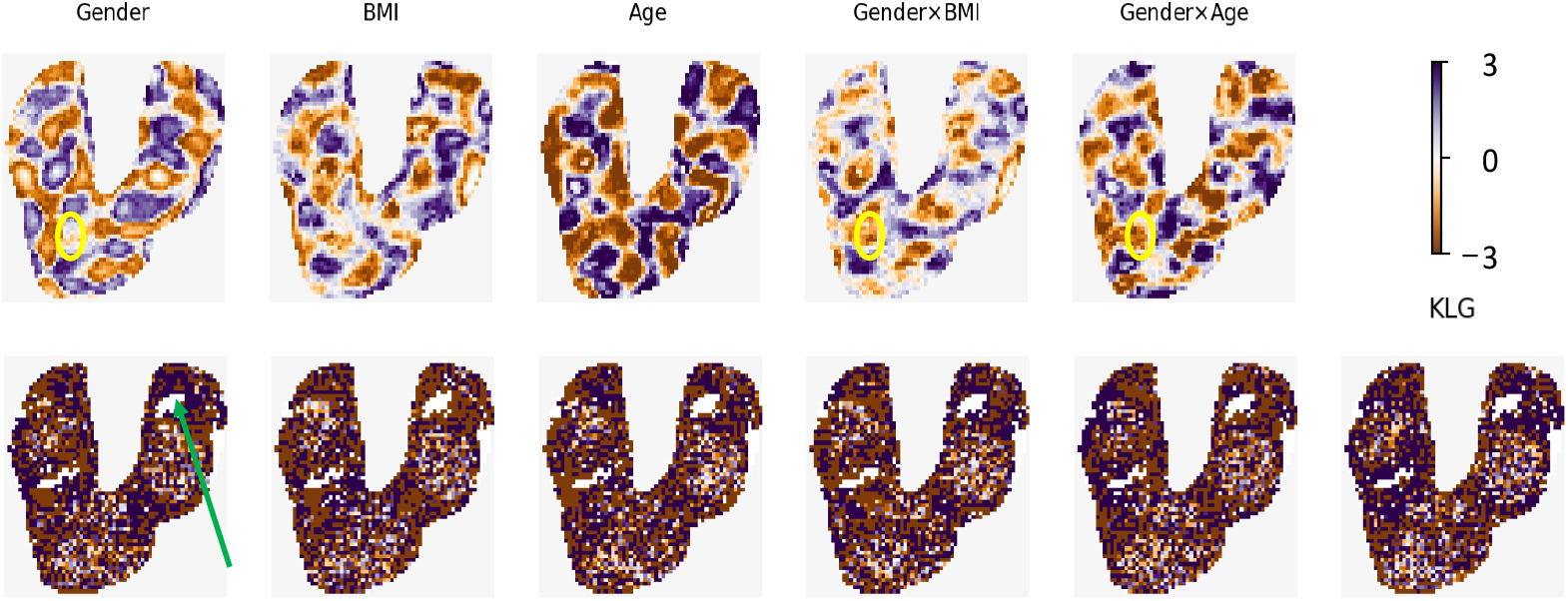
OAI data analysis: *t* test statistic maps of estimated ***α***_*s*_ in model (5) (top) and *t* test statistic maps of estimated ***β***_*s*_ and *φ*_*s*_ in model (7) (bottom).

The key findings are summarized as follows. First, with respect to overall average cartilage thickness, we observe high absolute *t*-statistics across multiple subregions of the left knee FC, reflecting the impact of various confounding factors. Notably, gender-related effects are not uniformly distributed. For example, in the subregion highlighted by the yellow circle in Figure 3, there is no significant difference between males and females. In contrast, BMI and Age exhibit more consistent and widespread associations with cartilage thickness throughout the left knee FC. Interestingly, interaction terms such as Gender × BMI and Gender × Age show significant effects in the same subregion, suggesting that older females with higher BMI are more prone to reduced cartilage thickness and smaller FC volumes. These findings align well with existing clinical evidence in the literature (Silverwood et al. 2015, Szilagyi et al. 2023). Second, different confounders appear to drive consistent spatial patterns of disease associations across most subregions of the left FC. For instance, certain regions, indicated by the green arrow in the second row of Figure 3, show no significant association with any of the covariates considered, indicating potential robustness or distinct biological behavior in those areas.

Next, to address the scientific question (Q3), i.e., the spatial location heterogeneity across patients, we investigated the detected disease regions and their heterogeneity across subjects. We randomly selected one OA patient from each of the four KLG groups (KLG=1,2,3, and 4), and their 2D FC thickness maps along with the detected disease regions are presented in Figure 4 (Columns 1 and 2). It can be found that the detected disease regions vary across subjects in terms of their number, size, shape, and location, which is consistent with our assumption of spatial heterogeneity. Besides the disease region at the individual level, we are also interested in the disease pattern at the population level. First, the PDPMs for each KLG-based subpopulation are presented in Figure 5. It can be found that the population-level disease pattern becomes more prevalent as the KLG level increases. In addition to the KLG-based population-level PDPM, we also derived the PDPMs for different potential subgroups with distinct disease patterns. Following the idea in Huang et al. (2022), we adopted the non-negative matrix factorization (NMF) method and applied it to the posterior probabilities of disease regions, 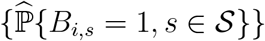, for all OA patients. We detected three subgroups, where the demographic information and the PDPM for each subgroup are presented in Table 2 and Figure 6(a), respectively.

**Table 2:** OAI data analysis: the detailed demographic information (Gender, BMI, Age, KLG) in each of the three detected subgroups. The mean and standard deviations for BMI and Age are reported for each subgroup as well.

|  | Gender (F/M) | BMI (kg/m<sup>2</sup>) | Age (years) | KLG (1/2/3/4) | Total |
| --- | --- | --- | --- | --- | --- |
| Subgroup 1 | 434/251 | 29.5±4.8 | 62.8±8.9 | 289/339/128/9 | 685 |
| Subgroup 2 | 266/250 | 29.9±4.5 | 62.7±8.7 | 83/132/211/90 | 516 |
| Subgroup 3 | 873/592 | 29.3±4.9 | 61.9±9.1 | 501/649/274/41 | 1465 |

**Figure 4:**
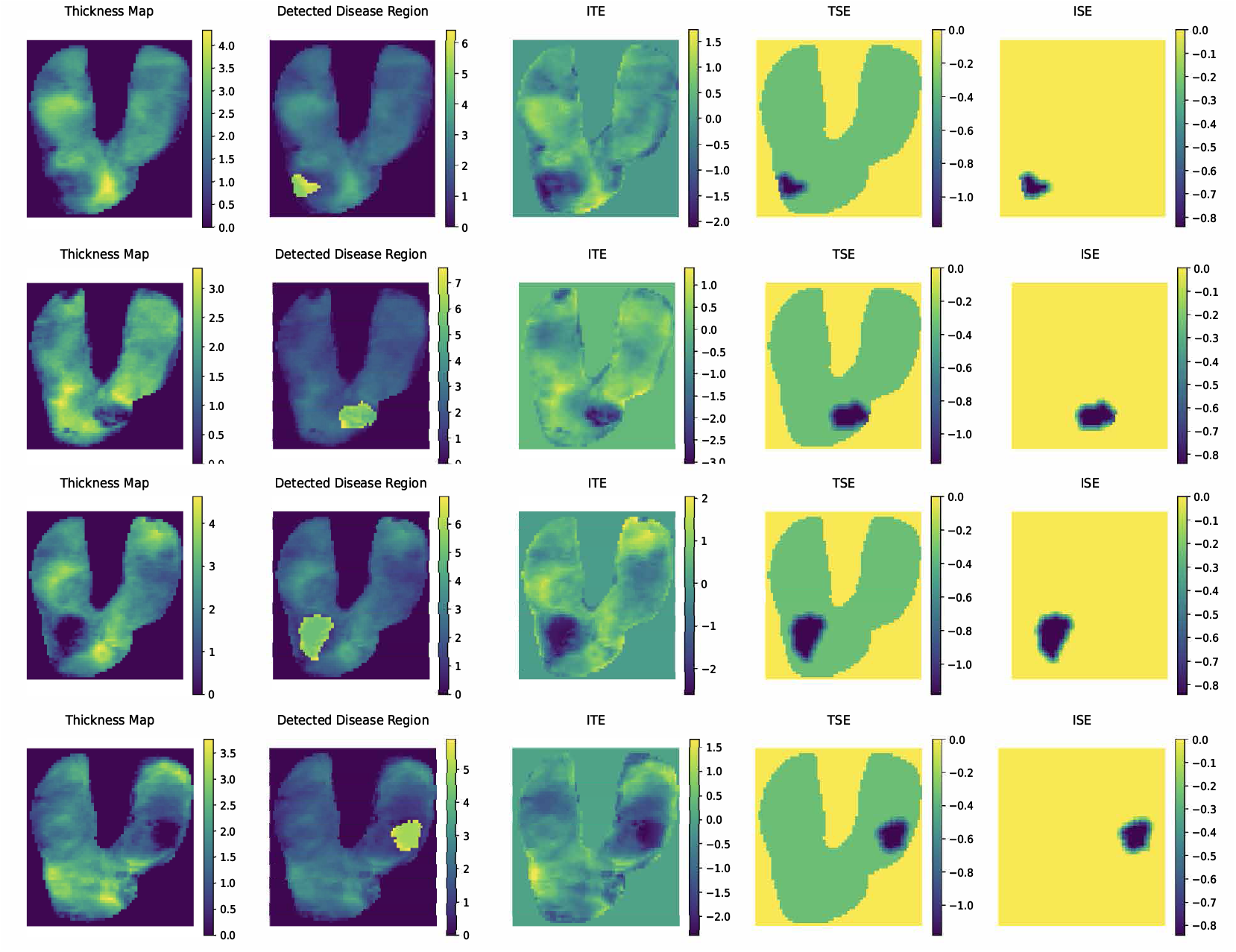
OAI data analysis: detected disease regions and estimated causal estimands for four patients randomly selected from different KLG groups (KLG=1,2,3,4 for row=1,2,3,4). Column 1: thickness map; Column 2: detected disease region; Column 3: estimated ITE; Column 4: estimated TSE; and Column 5: estimated ISE.

**Figure 5:**
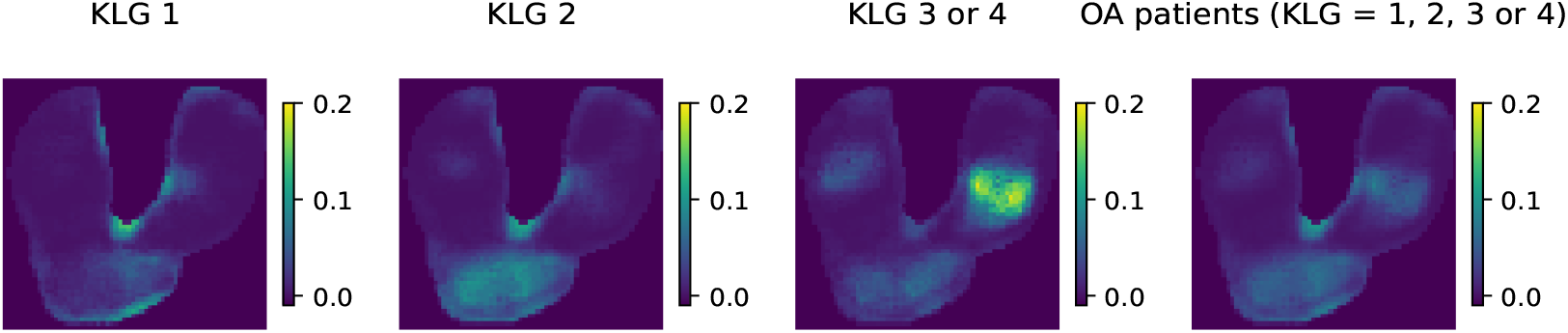
OAI data analysis: PDPM for different KLG-based subpopulations. Column 1: PDPM for patients with KLG=1; Column 2: PDPM for patients with KLG=2; Column 3: PDPM for patients with KLG=3 or 4; Column 4: PDPM for all OA patients (KLG=1,2,3 or 4).

**Figure 6:**
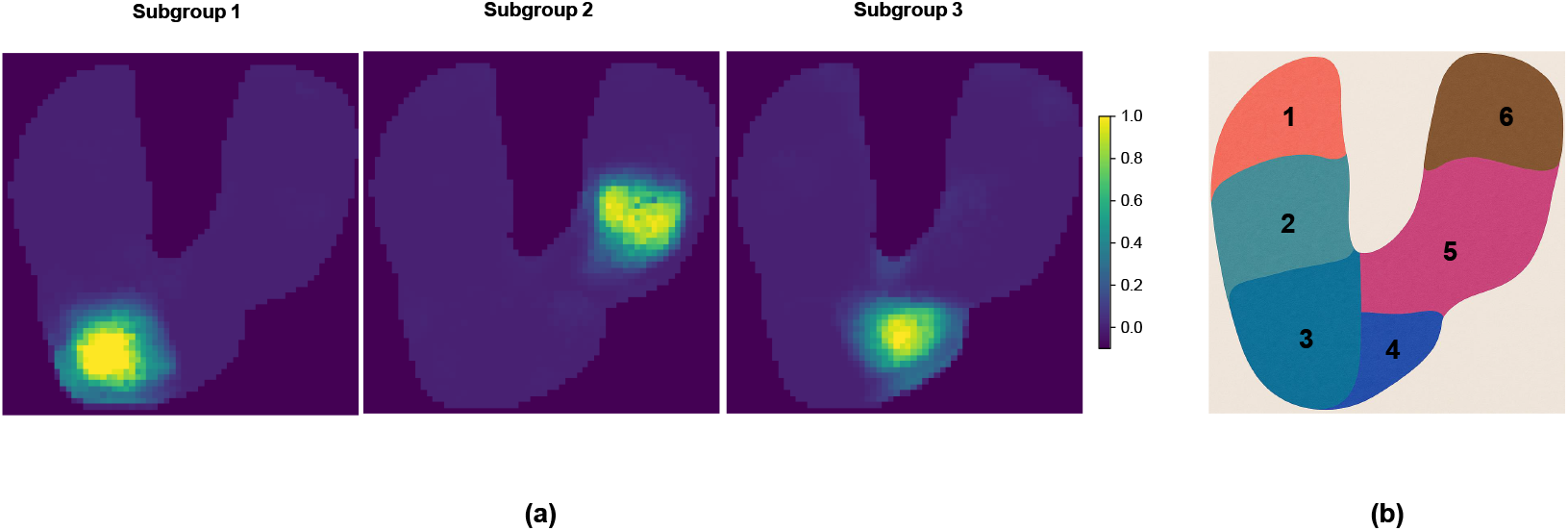
OAI data analysis: (a) PDPMs for three detected latent subgroups; (b) Parcellations of the left knee FC (Li, Luo, Chen, Huang, Shen, Xu et al. 2023): 1. Lateral Posterior Femur (LPF), 2: Lateral Central Femur (LCF), 3: Lateral Anterior Femur (LAF), 4: Medial Anterior Femur (MAF), 5: Medial Central Femur (MCF), and 6: Medial Posterior Femur (MPF).

Some key findings are summarized below. First, the three detected subgroups possess distinct disease patterns. According to the parcellations of the left knee FC (Li, Luo, Chen, Huang, Shen, Xu et al. 2023), the population disease patterns located in three different subregions (see Figure 6(b)), including Lateral Anterior Femur (LAF), Medial Central Femur (MCF), and Medial Anterior Femur (MAF). The cartilage thickness and volume in all of these subregions have been found commonly reduced among OA patients in existing literature (Neogi et al. 2009, Hayashi et al. 2014, Roemer et al. 2022). In particular, in one study focusing on the heterogeneity of cartilage damage (Roemer et al. 2022), most mild and severe OA patients had cartilage damage in MCF, which is consistent with the derived PDPM from Subgroup 2, in which 84% patients were assessed with KLG≥ 2. Furthermore, compared to Subgroups 1 and 3, Subgroup 2 involves patients with higher BMI and higher pecentage of severe OA patients (KLG=3 or 4), which is consistent with the disease pattern in the PDPM for patients with KLG=3 or 4 (Figure 5). Similar disease patterns have also been revealed in existing literature (Huang et al. 2022, Li, Luo, Chen, Huang, Shen, Xu et al. 2023).

To address the scientific questions (Q1), we calculated the estimated ITEs for all OA patients. The ITEs for selected OA patients are presented in Figure 4 (Column 3). It can be found that, the ITEs vary across patients and the OA-related effect (ITE with negative values) is highly consistent with the detected disease pattern. To further address the scientific questions (Q2), i.e., the causal pathway *g*_*i*_ → *B*_*i*_ → *Y*_*i*_, we fist estimated the KLG-related effect, 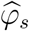, which is shown in Figure S6, and the corresponding *t*-statistic map shown in Figure 3. Then, we calculated the estimated causal estimands, including TSE, ISE, and DSE, for all OA patients. The TSE and ISE for the four randomly selected OA patients are presented in Figure 4 (Columns 4 and 5), and the estimated DSE is -0.21, which is invariant across pixels and subjects. The results demonstrate that the causal estimands strongly depend on the heterogeneous disease regions, and the spillover effects exist in the spatial causal effects across different patients, highlighting both spatial location heterogeneity between patients and spatial spillover effects within each subject.

Finally, similar to the simulation studies, we conducted two sensitivity analyses to evaluate robustness: (i) varying the threshold *p*_0_ in MCMC sampling and (ii) modifying Assumption 3.2 about unmeasured confounding. We also checked whether Assumptions 3.1 and 3.3 are reasonably satisfied in our real data analysis. In the first sensitivity analysis, we tested *p*_0_ at four different values, i.e., *p*_0_ = 0.3, 0.4, 0.6 and 0.7. Since there was no ground truth in our real data analysis, we set the estimated causal estimands with *p*_0_ = 0.5 as the gold standard. Then, under each setting of *p*_0_, posterior estimates of model parameters were obtained, and the MSEs of both TSE and ISE were calculated across diseased subjects. Results are summarized in Figure S7 (left). We observed that the ISE estimates remained stable across all *p*_0_ settings, whereas the MSE of the TSE increased as *p*_0_ grew larger. Given the relationship among TSE, ISE, and DSE, this result suggests that increasing *p*_0_ leads to smaller detected disease regions, which in turn adversely affects the accuracy of DSE estimation, i.e., estimation of *γ*. In the second sensitivity analysis, we assessed the impact of unmeasured confounding by testing *ξ*_*y*_ at three different values, i.e., *ξ*_*y*_ ∈ {0, 0.1, 0.2}, where *ξ*_*y*_ = 0 corresponds to the assumption of no unmeasured confounding. Thickness maps were simulated under model (8), using the previously estimated parameters from the real data as ground truth. We then refitted the proposed model to each simulated dataset and computed MSEs for the TSE and ISE across all subjects, treating the estimated TSE and ISE based on the original real data as the ground truth. The results, displayed in Figure S7 (right), indicate that the influence of unmeasured confounding becomes negligible when *ξ*_*y*_ ≥ 0.2, suggesting that the proposed method is robust even under mild violations of Assumption 3.2. Next, to assess Assumption 3.1, which requires the consistency of pixel-wise cartilage thickness distributions between normal controls and non-diseased regions in OA patients, we estimated the density functions using pixel-level residuals from two sources: (i) all pixels in the normal control group and (ii) pixels located in radiographically normal regions within each KLG-based subpopulation. Estimated density functions of these residuals, shown in Figure S8, reveal substantial overlap across all KLG-based subpopulations and the control group. To verify Assumption 3.3, which posits similar distributions of covariates between normal controls and OA patients, we compared the distributions of interaction terms, i.e., Gender*×*Age and Gender*×*BMI, for normal controls and different KLG-based subpopulations. Estimated density functions for these covariates, shown in Figures S9 and S10, exhibit consistent patterns across all subpopulations.

## 6 Conclusion

We have proposed a novel causal inference framework, HCDPD, designed to uncover complex causal pathways linking early-stage diseases to latent disease patterns and their manifestations in organs, as observed through later-stage medical imaging. By leveraging advanced Bayesian inference techniques, our method accurately estimates both direct and indirect causal effects within the HCDPD framework. We have validated our approach using the OAI dataset, where it has successfully identified and characterized diverse disease patterns across different patients. Notably, this innovative framework is versatile and can be adapted to analyze imaging datasets for various other diseases.

Despite these advancements, several avenues for future research remain. First, it would be valuable to investigate how the clinical diagnosis *g*_*i*_ and baseline covariates **x**_*i*_ influence the size of the disease region. Given that 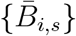 can be interpreted as the relative size of the local disease region, we could develop a deep regression model (DRM) to examine the relationship between {*g*_*i*_, **x**_*i*_} and 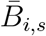 across pixels. To achieve this, we could identify a subset of pixels, 𝒮_0_ ⊂ 𝒮, such that their corresponding neighborhoods {𝒩 (*s*), *s* ∈ 𝒮_0_} form a partition of 𝒮. The proposed DRM can then be formulated as: 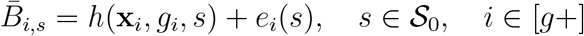, where *h*(·) is an unknown function approximated by a deep learning architecture, such as a multilayer perceptron (MLP). Additionally, since the OAI study is a longitudinal cohort, it would be highly beneficial to extend our causal framework to a spatiotemporal version. This extension would allow for the comprehensive modeling of disease pattern heterogeneity across subjects, pixels, and time points, significantly enhancing the framework’s applicability in longitudinal studies.

## Data Availability

Github

## Acknowledgment

The authors used ChatGPT solely for language editing and proofreading.

## Notes

Drs. Niethammer and Zhu are partially supported by the National Institutes of Health (NIH) grants 1R01AR082684 and 1OT2OD038045-01. Dr. Zhu is also partially supported by the Gillings Innovation Laboratory on gen- erative AI and the National Institute on Aging (NIA) of the National Institutes of Health (NIH) grants U01AG079847, 1R01AG085581, RF1AG082938, and R01AR082684. The content is solely the responsibility of the authors and does not necessarily represent the official views of these institutes.

### Competing Interest Statement

The authors have declared no competing interest.

### Funding Statement

National Institutes of Health (NIH) grants 1R01AR082684 and 1OT2OD038045-01. National Institutes of Health (NIH) grants U01AG079847, 1R01AG085581, RF1AG082938, and R01AR082684.

